# Infection with the SARS-CoV-2 Delta Variant is Associated with Higher Infectious Virus Loads Compared to the Alpha Variant in both Unvaccinated and Vaccinated Individuals

**DOI:** 10.1101/2021.08.15.21262077

**Authors:** Chun Huai Luo, C. Paul Morris, Jaiprasath Sachithanandham, Adannaya Amadi, David Gaston, Maggie Li, Nicholas J. Swanson, Matthew Schwartz, Eili Y. Klein, Andrew Pekosz, Heba H. Mostafa

## Abstract

**Background:** The emerging SARS-CoV-2 variant of concern (VOC) B.1.6.17.2 (Delta) quickly displaced the B.1.1.7 (Alpha) and is associated with increases in COVID-19 cases nationally. The Delta variant has been associated with greater transmissibility and higher viral RNA loads in both unvaccinated and fully vaccinated individuals. Data is lacking regarding the infectious virus load in Delta infected individuals and how that compares to individuals infected with other SARS-CoV-2 lineages.

**Methods:** Whole genome sequencing of 2,785 clinical isolates was used to characterize the prevalence of SARS-CoV-2 lineages circulating in the National Capital Region between January and July 2021. Clinical chart reviews were performed for the Delta, Alpha, and B.1.2 (a control predominant lineage prior to both VOCs) variants to evaluate disease severity and outcome and Cycle threshold values (Cts) were compared. The presence of infectious virus was determined using Vero-TMPRSS2 cells and anti-SARS-CoV-2 IgG levels were determined from upper respiratory specimen. An analysis of infection in unvaccinated and fully vaccinated populations was performed.

**Results:** The Delta variant displaced the Alpha variant to constitute 88.2% of the circulating lineages in the National Capital Region by July, 2021. The Delta variant associated with increased breakthrough infections in fully vaccinated individuals that were mostly symptomatic when compared to the Alpha breakthrough infections, though it is important to note there was a significantly longer period of time between vaccination and infection with Delta infections. The recovery of infectious virus on cell culture was significantly higher with the Delta variant compared to Alpha in both vaccinated and unvaccinated groups. The impact of vaccination on reducing the recovery of infectious virus from clinical samples was only observed with Alpha variant infections but was strongly associated with low localized SARS-CoV-2 IgG for both variants. A comparison of Ct values showed a significant decrease in the Delta compared to Alpha with no significant differences between unvaccinated and vaccinated groups.

**Conclusions:** Our data indicate that the Delta variant is associated with increased infectious virus loads when compared to the Alpha variant and decreased upper respiratory antiviral IgG levels. Measures to reduce transmission in addition to increasing vaccinations rates have to be implemented to reduce Delta variant spread.

**Funding:** NIH/NIAID Center of Excellence in Influenza Research and Surveillance contract HHS N2772201400007C, Johns Hopkins University, Maryland department of health, Centers for Disease Control and Prevention contract 75D30121C11061.

## Introduction

SARS-CoV-2 genomic evolution triggered concerns for the emergence of variants that could be more transmissible, cause severe disease, or escape natural or vaccine induced protective immunity. Lineage B.1.1.7 (Alpha) was classified as a variant of concern (VOC) by the United States Center for Disease Control and Prevention (CDC) due to evidence of higher transmissibility and concern for more severe disease (1). The Alpha variant, which was first detected in Southeast England in September 2020 (2), became the predominant in new SARS-CoV-2 infections in the UK by December 2020, spread globally, and rapidly became the major lineage in the US by April 2021 (3, 4). Estimates from the UK suggested the Alpha variant was 50% more transmissible and had a 43-90% higher reproduction number compared to other SARS-CoV-2 lineages (5). Although early reports found no correlation between Alpha and increased severity of disease (5), other studies reported an association with higher mortality (6) and risk of hospitalization (7).

The B.1.617.2 (Delta) variant displaced the Alpha in the United States after a nationwide decline in the total numbers of cases in June 2021 and became the most frequently sequenced lineage by July 2021 (8). The Delta variant was classified as a VOC by the WHO in May 2021 due to a notable increased transmissibility, even in the context of increasing percentages of fully vaccinated individuals in various communities. The Delta variant was associated with SARS-CoV-2 outbreaks and breakthrough infections in vaccinated individuals (9, 10), however, while vaccination is most effective against preventing severe disease, subsequent hospitalization, and death from the Delta variant, it appears to be less effective against infection and perhaps even transmission (11-13).

Greater understanding of the reasons for increased transmissibility in both the Alpha and Delta variants is urgently needed to understand the potential for breakthrough infections and outbreaks in the Fall and Winter. One proposed hypothesis is that the variants are able to attain higher viral loads in the respiratory tract of infected individuals (5, 14). While some reports have found an association between the Alpha and Delta variants and higher viral loads in the upper respiratory tract (5, 6, 10), greater understanding is needed of the relative replicability of these variants. Quantifying these differences is particularly important in vaccinated individuals, where the Delta variant has been associated with comparable Ct values in vaccinated versus unvaccinated patients (15). Higher replicability could additionally be important in vaccinated individuals with waning immunity. A few studies have assessed the recovery of infectious virus and neutralizing antibodies (16, 17). In this study, we used a large cohort of samples characterized by whole genome sequencing between January 2021 and July 2021 to compare the clinical characteristics, Ct values from upper respiratory specimens, recovery of infectious virus, and nasal SARS-CoV-2 IgG for Delta and Alpha variants.

## Methods

### Ethical considerations and Data availability

The research Johns Hopkins Medical Institutions Institutional Review Board-X (JHM IRB-X) is constituted to meet the requirements of the Privacy Rule at section 45 CFR 164.512(i)(1)(i)(B) and is authorized and qualified to serve as the Privacy Board for human subjects research applications conducted by Johns Hopkins University faculty members. JHM IRB-3 approved IRB00221396 entitled “Genomic evolution of viral pathogens: impact on clinical severity and molecular diagnosis”. IRB review included the granting of a waiver of consent based on the following criteria: 1) the research involves no more than minimal risk to subjects; 2) the waiver will not adversely affect the rights and welfare of the subjects; 3) the research could not be practicably carried out without the waiver; and 4) the IRB will advise if it is appropriate for participants to be provided with additional pertinent information after participation. This study was also approved for the inclusion of children as ‘research not involving greater than minimal risk’. The permission of parents/guardians is waived. Assent is waived for all children. JHM IRB-X determined that there is no requirement for continuing review or progress report for this application. Remnant nasopharyngeal or lateral mid-turbinate nasal (NMT) clinical swab specimens from patients who tested positive for SARS-CoV-2 after the standard of care testing were used.

### Specimens and Patient Data

The clinical specimens used for sequencing were nasopharyngeal or lateral mid-turbinate nasal swabs after standard of care diagnostic or screening assays were performed during inpatient and outpatient encounters across the Johns Hopkins Medical System, which encompasses five acute care hospitals and more than 40 ambulatory care offices. In addition, specimens were obtained through standard of care screening and testing services performed by the health system at several long-term care facilities in the State of Maryland as well as through mobile outreach clinics in local neighborhoods. Molecular assays used for diagnosis include RealStar® SARS-CoV-2 RT-PCR (Altona Diagnostics), Xpert Xpress SARS-CoV-2/Flu/RSV (Cepheid), NeuMoDx SARS-CoV-2 (Qiagen), Cobas SARS-CoV-2 (Roche), ePlex Respiratory Pathogen Panel 2 (Roche), Aptima SARS-CoV-2 (Hologic), and Accula SARS-CoV-2 assays (ThermoFisher Scientific) (18-21). Molecular diagnosis of SARS-CoV-2 at Johns Hopkins Hospital laboratory began on March 11 2020 (22), and whole genome sequencing for identifying circulating SARS-CoV-2 variants started as early as March 2020 as well (22). Surveillance efforts for VOCs were increased at the end of October 2020 to monitor the evolution of SARS-CoV-2. Unique patients’ specimens were used for this study.

### Ct value analyses

Samples with available Ct values after clinical diagnosis was made with NeuMoDx SARS-CoV-2 (Qiagen) testing were included in this study (N gene), as were samples retested after initial diagnostic testing using the PerkinElmer® New Coronavirus Nucleic Acid Detection Kit (https://www.fda.gov/media/147547/download) to ensure comparable Ct values (N gene).

### Amplicon based Sequencing

Specimens were extracted using the Chemagic™ 360 system (Perkin Elmer) following the manufacturer’s protocol. 300 µL of sample was extracted and eluted in 60 µL elution buffer. Sequencing and data analysis were performed as previously described (4, 22).

### Cell culture

Vero-TMPRSS2 cells were cultured and infected with aliquots of swab specimens as previously described for VeroE6 cells (23). The presence of SARS-CoV-2 was confirmed by reverse transcriptase PCR (qPCR).

### ELISA

Undiluted respiratory samples were tested with the EUROIMMUN Anti-SARS-CoV-2 ELISA (IgG) following the package insert (https://www.fda.gov/media/137609/download). The assay detects antibodies to the S1 domain of the spike protein of SARS-CoV-2 with a cut-off < 0.8 for negative results and ≥ 0.8 to < 1.1 as borderline. The 1.1 value was used as a cut off for respiratory specimen types.

### Clinical data analysis

Clinical data were collected by retrospective review of electronic medical records. Events leading to hospitalization, requirement of ICU level care, and mortality were evaluated and only patients with these events clearly attributable to COVID-19 were included in these analyses. Vaccine breakthrough infections were based on the CDC definition of with positive test results more than 14 days post the second shot for pfizer/BioNTech BNT162b2 and Moderna mRNA-1273 or 14 days after the J&J/ Janssen shot.

### Statistical analysis

Statistical analyses were conducted using GraphPad prism. Chi-square and Fisher Exact tests were used for categorical variable comparisons and t-test and Kruskal-Wallis one-way ANOVA tests were used for comparing continuous independent variables.

## Results

### SARS-CoV-2 Positivity and Variants trends

Between January 2021 and July 2021, a total of 209,905 samples were tested at the Johns Hopkins Hospital Laboratory with positivity rates that declined from 7.7% in January to 0.7% in June with a slower increase noted in July (Figure 1A). Of 2,785 genomes sequenced in this time frame, our data showed that the predominant circulating lineages (primarily B.1.2, clade 20G) were displaced by Alpha in late February (4) which was subsequently displaced by Delta at the end of June (Figure 1B). Other VOC and VOI were detected only infrequently during this time frame (Figure 1B).

**Figure 1.**
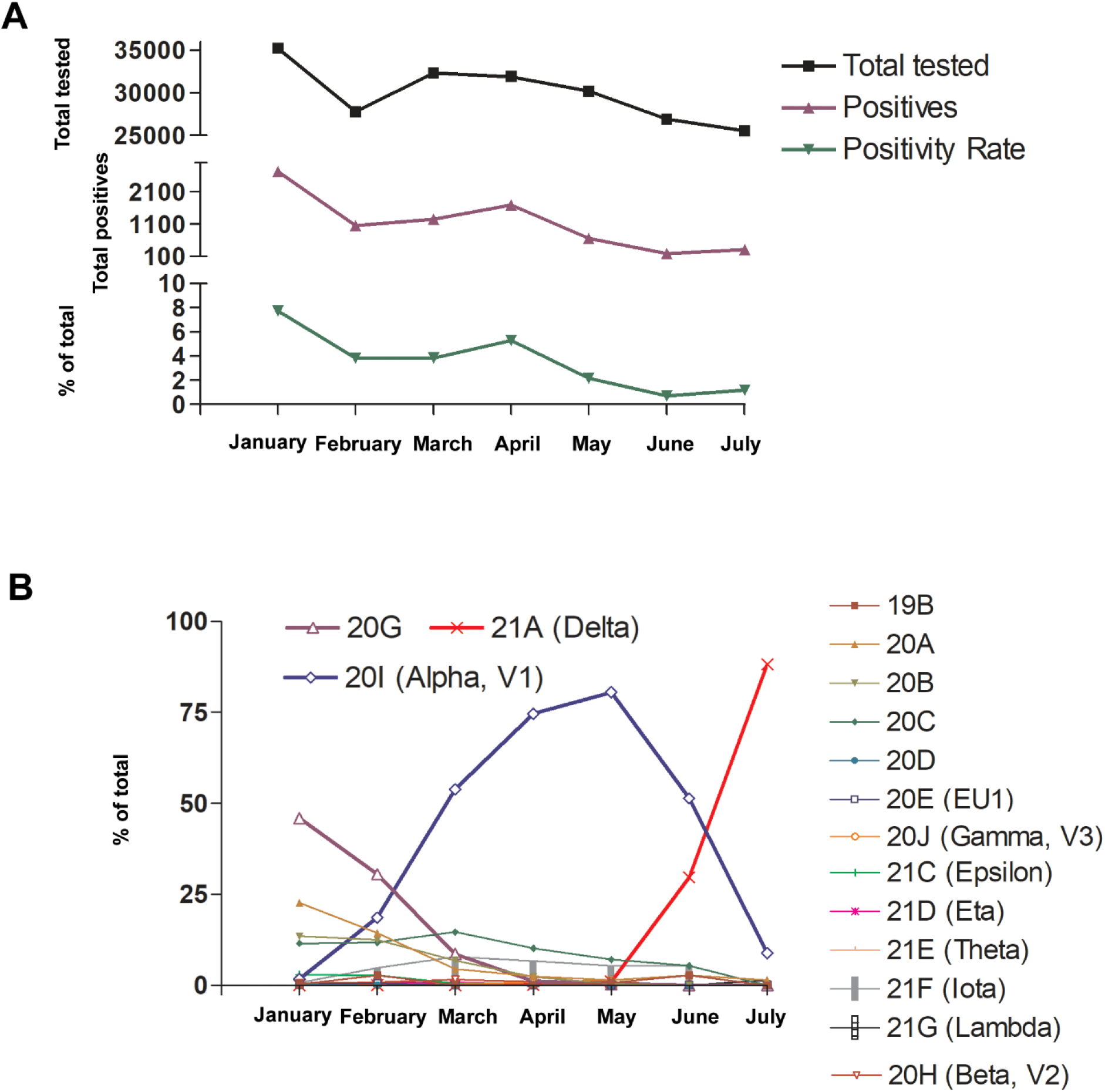
A) SARS-CoV-2 molecular testing at Johns Hopkins Laboratory showing total tested, total positives, and % positivity from January to July 2021 B) Circulating SARS-CoV-2 clades including VOC and VOI from January to July 2021.

### Patient characteristics and infection outcomes in Alpha and Delta infections

Clinical chart reviews were performed for all Delta (107), Alpha (1482), and B.1.2 (377) infected patients diagnosed at Johns Hopkins laboratory from January to July 2021. The Delta variant caused a significant increase in confirmed breakthrough infections when compared to the Alpha variant (28% vs 12.4%, p < 0.00001, Table 1). Not surprisingly, there was a significant increase in the median days after vaccination for the Delta variant breakthroughs compared to Alpha variant breakthroughs (136.3 vs 20.1, p < 0.00001, Table 1) which reflects the lack of Delta variants in our geographic region during the initial COVID-19 vaccine rollout. Delta variant infected patients were significantly different in race distribution when compared to the Alpha with a significant increase in infections in white race (p = 0.005), however, African-American patients predominated infections with both variants. No differences in hospital admission or mortality were noted between the Delta and Alpha variants however, the Delta infected group had a lower prevalence of certain comorbidities including kidney disease and diabetes (Table 1).

**Table 1.**
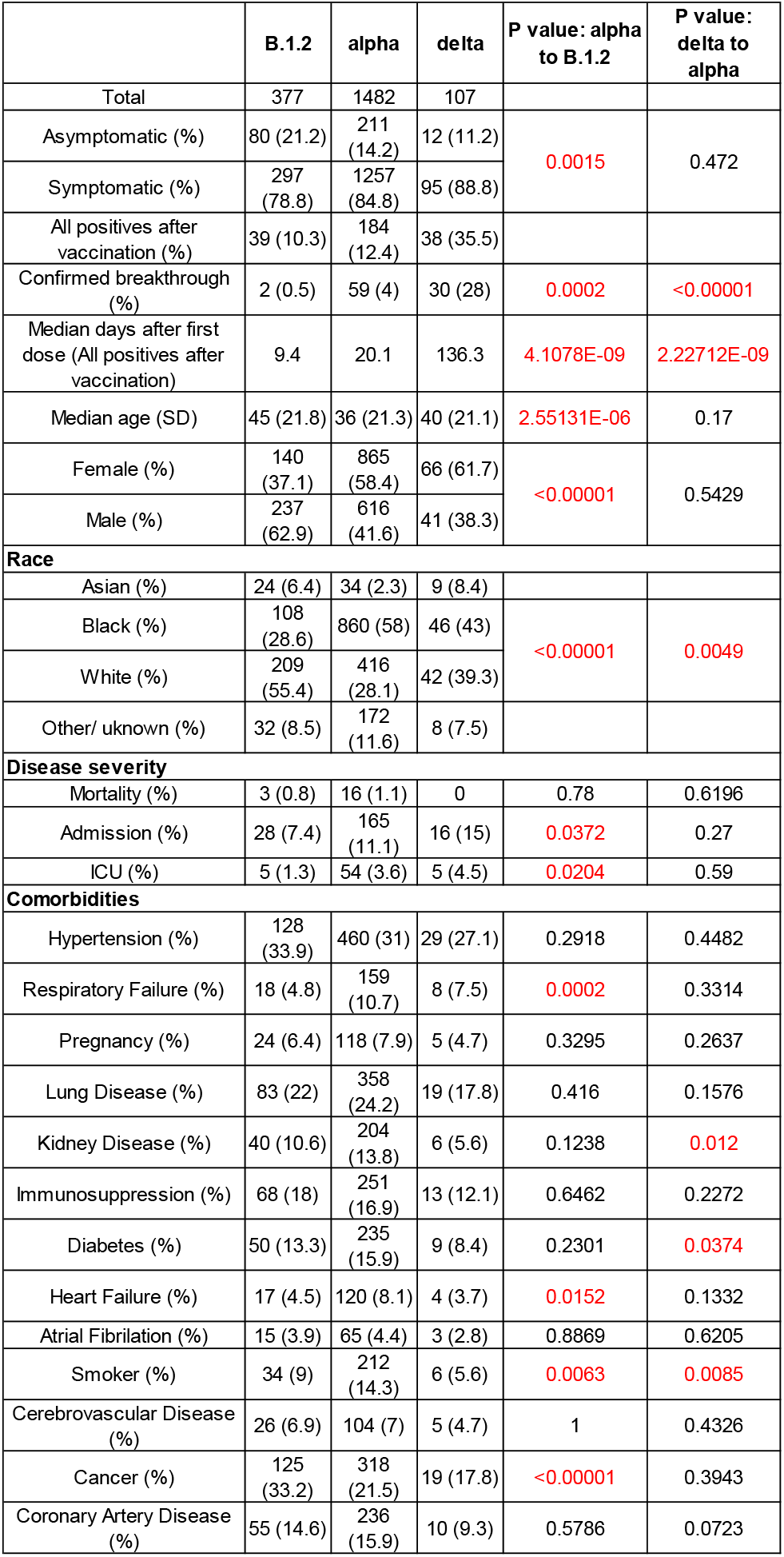
Clinical and metadata of the B.1.2, Alpha, and Delta infected patients. Statistics for ages and median days after vaccination were calculated by t test and all other statistics were calculated by Chi-squared test.

The Alpha variant was associated with a significant increase in symptomatic infections when compared to the precedent B.1.2 lineage (84.8% vs 78.8, p = 0.0015, Table 1). However, there was no similar increase from Alpha to Delta (p=0.472, Table 1). A reduction in the median age, reduction in the male to female ratio and an increase in infections in the self-identify as African-American were noted for the Alpha variant compared to the B.1.2 lineage (Table 1). When compared to the B.1.2, the Alpha variant showed a significant increase in COVID related hospitalization and ICU level care, but not mortality.

When vaccine breakthrough infection cases were compared to the unvaccinated patients in the Alpha and Delta groups, no significant differences in the likelihood of COVID related hospital admissions were observed, however the Alpha and to a lesser extent the Delta vaccine breakthrough groups were characterized by significantly higher immunosuppression and other comorbidities (Table 2). Comorbidities including hypertension, kidney disease, heart failure, and coronary artery disease were associated with vaccine breakthrough infections with the Alpha, but not the Delta variants (Table 2). Vaccine breakthrough infections with the Delta variant were associated with a significantly higher percentage of symptomatic infections (93.3% vs 61%, p = 0.001), reduced median age (40.5 years vs 51 years, p = 0.035), and a marked increase in the median days after receiving the vaccine (152.7 days vs 77.3 days, p < 0.00001, Table 2).

**Table 2.** Clinical and metadata of Delta and Alpha vaccinated and unvaccinated patients. All positives after vaccination includes any patient who received vaccination prior to the positive test result. True breakthrough infections were based on the CDC definition to include positives more than 14 days after the second dose for pfizer/BioNTech BNT162b2 or Moderna mRNA-1273 or 14 days after the J&J/ Janssen shot. Statistics for ages and median days after vaccination were calculated by t test and all other statistics were calculated by Chi-squared test.

|  | Delta |  |  |  |  | Alpha |  |  |  |  | P value/ alpha to delta true breakthrough |
| --- | --- | --- | --- | --- | --- | --- | --- | --- | --- | --- | --- |
|  | All positives after vaccination | True breakthrough | Unvaccinated | P value: all positives after vaccination to unvaccinated | P value: true breakthrough to unvaccinated | All positives after vaccination | True breakthrough | Unvaccinated | P value: all positives after vaccination to unvaccinated | P value: true breakthrough to unvaccinated |  |
| Total | 38 | 30 | 69 |  |  | 184 | 59 | 1298 |  |  |  |
| Median days after 1st dose (SD) |  | 152.7 (59.8) |  |  |  |  | 77.3 (27.9) |  |  |  | 3.511E-06 |
| Symptomatic (%) | 34 (89.5) | 28 (93.3) | 61 (88.4) | 1 | 0.72 | 148 (80.4) | 36 (61) | 1109 (85.4) | 0.0424 | <0.00001 | 0.0011 |
| Asymptomatic (%) | 4 (10.5) | 2 (6.7) | 8 (11.6) |  |  | 36 (19.6) | 23 (38.9) | 175 (13.5) |  |  |  |
| Median age | 40.5 | 40.5 | 37 | 0.024 | 0.039 | 53 | 51 | 34 | 2.24559E-26 | 0.142357054 | 0.035 |
| Females (%) | 22 (57.9) | 18 (60) | 44 (63.8) | 0.67 | 0.82 | 112 (60.9) | 42 (71.2) | 753 (58) | 0.52 | 0.06 | 0.34 |
| Males (%) | 16 (42.1) | 12 (40) | 25 (36.2) |  |  | 72 (39.1) | 17 (28.8) | 544 (41.9) |  |  |  |
| Race |  |  |  |  |  |  |  |  |  |  |  |
| Asian (%) | 5 (13.2) | 5 (16.7) | 4 (5.8) |  |  | 4 (2.2) | 1 (1.7) | 30 (2.3) |  |  |  |
| Black (%) | 11 (28.9) | 6 (20) | 35 (50.7) |  |  | 73 (39.7) | 13 (22) | 787 (60.6) |  |  |  |
| White (%) | 20 (52.6) | 18 (60) | 22 (31.9) | 0.026 | 0.0035 | 86 (46.7) | 38 (64.4) | 330 (25.4) | <0.00001 | <0.00001 | 1 |
| Other/ unknown (%) | 2 (5.3) | 1 (3.3) | 8 (11.6) |  |  | 21 (11.4) | 7 (11.9) | 151 (11.6) |  |  |  |
| Disease severity |  |  |  |  |  |  |  |  |  |  |  |
| Admitted (%) | 4 (10.5) | 3 (10) | 12 (17.4) | 0.4 | 0.54 | 26 (14.1) | 4 (6.8) | 139 (10.7) | 0.17 | 0.51 | 0.11 |
| ICU level care (%) | 3 (7.9) | 2 (6.7) | 2 (2.9) | 0.3445 | 0.58 | 8 (4.3) | 2 (3.4) | 46 (3.5) | 0.53 | 1 | 0.6 |
| Death (%) | 0 | 0 | 0 |  |  | 4 (2.2) | 0 | 12 (0.9) | 0.13 | 1 |  |
| Comorbidities |  |  |  |  |  |  |  |  |  |  |  |
| Hypertension (%) | 13 (34.2) | 10 (33.3) | 16 (23.2) | 0.26 | 0.33 | 91 (49.5) | 26 (44.1) | 369 (28.4) | <0.00001 | 0.0125 | 0.36 |
| Respiratory Failure (%) | 5 (13.2) | 4 (13.3) | 3 (4.3) | 0.129 | 0.19 | 24 (13) | 7 (11.9) | 135 (10.4) | 0.307 | 0.66 | 1 |
| Pregnancy (%) | 2 (5.3) | 2 (6.7) | 3 (4.3) | 1 | 0.64 | 13 (7.1) | 7 (11.9) | 105 (8.1) | 0.77 | 0.33 | 0.7 |
| Lung Disease (%) | 6 (15.8) | 4 (13.3) | 13 (18.8) | 0.79 | 0.77 | 47 (25.5) | 14 (23.7) | 311 (23.9) | 0.65 | 1 | 0.28 |
| Kidney Disease (%) | 4 (10.5) | 3 (10) | 2 (2.9) | 0.182 | 0.16 | 45 (24.5) | 13 (22) | 159 (12.2) | 0 | 0.042 | 0.24 |
| Immunosuppression (%) | 9 (23.7) | 6 (20) | 4 (5.8) | 0.011 | 0.06 | 53 (28.8) | 15 (25.4) | 198 (15.3) | 0 | 0.044 | 0.79 |
| Diabetes (%) | 5 (13.2) | 5 (16.7) | 4 (5.8) | 0.275 | 0.12 | 51 (27.7) | 13 (22) | 184 (14.2) | 0 | 0.127 | 0.78 |
| Heart Failure (%) | 1 (2.6) | 1 (3.3) | 3 (4.3) | 1 | 1 | 29 (15.8) | 10 (16.9) | 91 (7) | 0.0002 | 0.0096 | 0.09 |
| Atrial Fibrillation (%) | 2 (5.3) | 2 (6.7) | 1 (1.4) | 0.286 | 0.22 | 19 (10.3) | 5 (8.5) | 46 (3.5) | 0.0002 | 0.07 | 1 |
| Smoker (%) | 3 (7.9) | 2 (6.7) | 3 (4.3) | 0.66 | 0.63 | 25 (13.6) | 5 (8.5) | 187 (14.4) | 0.8227 | 0.25 | 1 |
| Cerebrovascular Disease (%) | 2 (5.3) | 1 (3.3) | 3 (4.3) | 1 | 1 | 26 (14.1) | 5 (8.5) | 78 (6) | 0.0003 | 0.40 | 0.65 |
| Cancer (%) | 12 (31.6) | 11 (36.7) | 7 (10.1) | 0.0081 | 0.0035 | 87 (47.3) | 31 (52.5) | 231 (17.8) | <0.00001 | <0.00001 | 0.18 |
| Coronary Artery Disease (%) | 3 (7.9) | 2 (6.7) | 7 (10.1) | 1 | 0.72 | 55 (29.9) | 18 (30.5) | 181 (13.9) | <0.00001 | 0.002 | 0.014 |

### Delta and Alpha variants cycle threshold values (Ct) in upper respiratory samples

To determine if the Ct values in respiratory specimens were different between Alpha, Delta, and B.1.2 variants, we compared the Ct values available for each group (N: B.1.2 = 224, Alpha = 562, Delta = 98) and associated the Ct values to the days after the onset of symptoms for symptomatic patients (N: B.1.2 = 214, Alpha = 511, Delta = 75). The mean Ct value for the Delta and B.1.2 variant was significantly lower when compared to the Alpha variant (20.08 vs 19.62 vs 21.74, p <0.05; Figure 2A). Similar trends were noted when Ct values were associated with samples collected within 5 days or less from symptoms onset (mean Ct for Alpha 20.98 vs 19.33 for Delta vs 19.25 for B.1.2, Figure 2B). For samples collected more than 5 days from symptoms, mean Ct values of the Alpha was significantly higher than the B.1.2 (24.4 vs 21.13, p < 0.05, Figure 2C).

**Figure 2.**
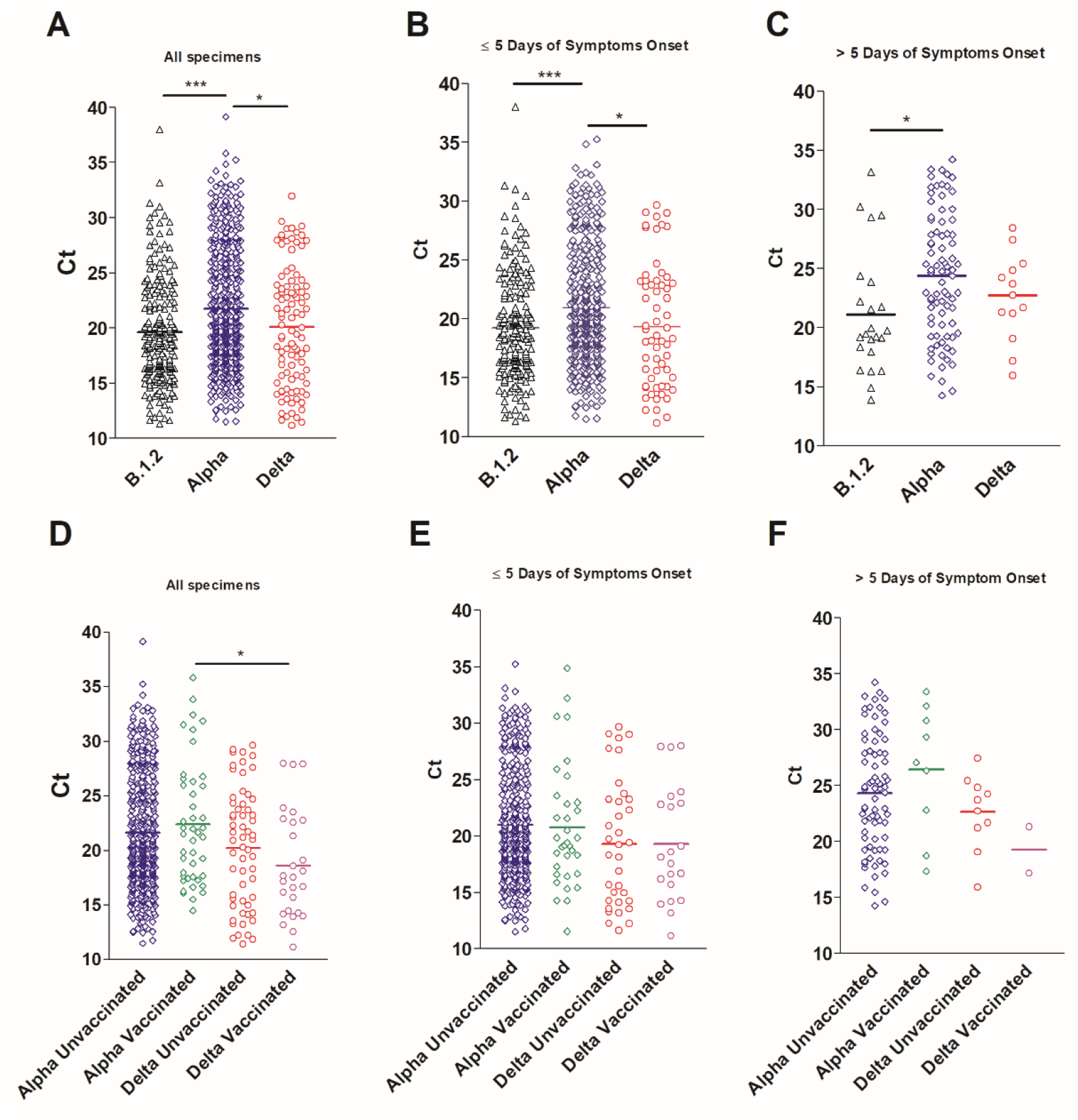
Cycle threshold (Ct) values of Alpha, Delta, and control variants. A) Ct values of B.1.2 (N = 254), Alpha (N = 562), and Delta (N = 98) variants from all samples with available Ct values. B and C) Correlation of Ct values and ranges of days after the onset of symptoms B) 0 to 5 days, C) >5 days. For this analysis, samples from asymptomatic patients were not included (N: B.1.2 = 214, Alpha = 511, Delta = 75). D) Ct values of Alpha and Delta variants broken down by vaccination status. Alpha (unvacccinated, N = 442, fully vaccinated N = 42), Delta (unvaccinated, N = 47, fully vaccinated N = 23). For this analysis, samples from partially vaccinated patients were not included. E and F) Correlation of Ct values and ranges of days after the onset of symptoms of the Alpha and Delta variants divided by the vaccination status. E) 0 to 5 days, F) >5 days. One-way ANOVA * p < 0.05, ** p < 0.001, ***, p < 0.0001.

Mean Ct values were significantly lower in Delta versus Alpha variant vaccine breakthrough groups (22.42 vs 18.6, p < 0.05) (Figure 2D), but no significant differences were observed between vaccinated and unvaccinated Ct values within each lineage. However, Alpha variant breakthrough vaccinated individuals demonstrated a noticeable change in mean Ct values when associated with the course of the infection (mean 20.75 within the first 5 days vs 26.45 after 5 days), but a similar analysis was not possible for the Delta variant breakthrough infections due to the infrequent positives after 5 days of symptoms in our cohort (Figure 2E and F)

### Recovery of infectious virus in Delta versus Alpha groups

To assess the recovery of infectious virus from Delta versus Alpha variant infected groups, samples from a total of 141 Alpha (95 from unvaccinated and 46 from vaccine breakthrough infections) and 90 Delta (63 unvaccinated and 27 from vaccine breakthrough infections) were used to inoculate Vero-TMPRSS2 cells. Significantly more specimens with Delta variants had infectious virus present as compared to specimens containing Alpha variants (Delta 67.8%, Alpha 31.2%; Figure 3A, p < 0.00001) Specimens from the fully vaccinated Alpha group showed significant reduction in the recovery of infectious virus as compared to the unvaccinated Alpha group (17.4% vs 37.9%, p = 0.02, Figure 3A) which was not the case in the Delta groups which had nearly equivalent specimens with infectious virus (70.4% vs 66.7%, Figure 3A). A significant increase in the recovery of infectious virus from specimens of patients infected with the Delta variant as compared to the Alpha variant was noted for both unvaccinated (66.7% vs 37.9%, p = 0.0006) and fully vaccinated (70.4% vs 17.4%, p < 0.00001) groups (Figure 3A). The mean Ct value for specimens associated with infectious virus (CPE) in all groups was significantly lower than groups without infectious virus (CPE positive: Delta unvaccinated, 17.6, Delta vaccinated, 16.1, Alpha unvaccinated, 18.1, Alpha vaccinated, 17.8-CPE negative: Delta unvaccinated, 25.3, Delta vaccinated, 24.4, Alpha unvaccinated, 24.9, Alpha vaccinated, 24.1, p < 0.0001) but no differences in mean Cts were noted between Alpha and Delta vaccinated and unvaccinated groups in infectious virus positive or negative groups (Figure 3B). Notably, the majority of the cell culture positives in the Alpha group showed CPE on day 5 after infecting the cellular monolayer (31.8%) in contrast to day 4 for the Delta group (63.9%), even though a few specimens in the Alpha group showed CPE as early as day 2 (15.9%) (Figure 3C).

**Figure 3.**
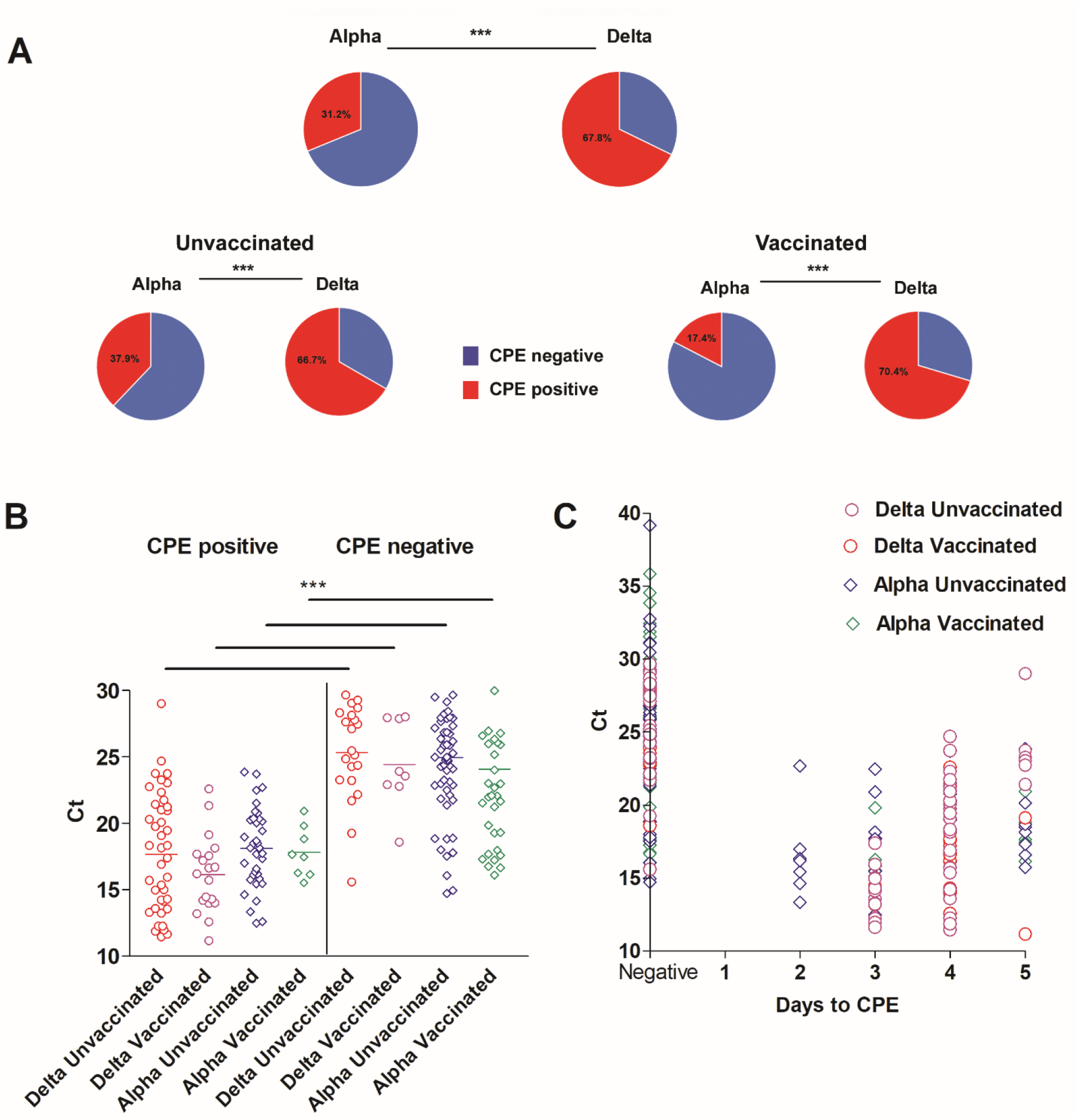
Recovery of infectious SARS-CoV-2 on Vero-TMPRSS2 cells for Alpha and Delta variants. A) Percent CPE positives and negatives for Alpha and Delta total, unvaccinated, and vaccinated groups (Alpha unvaccinated, N = 95, vaccinated N = 46, Delta unvaccinated, N = 63, vaccinated, N = 27). Chi-squared test *** p < 0.0001. B) Ct range of CPE positive and negative Alpha and Delta variants. One-way ANOVA *** p < 0.0001 C) Correlation of Ct values and days to the first appearance of CPE (cytopathic effect) for the Alpha and Delta groups.

### Localized SARS-CoV-2 IgG in the Delta versus the Alpha groups

To assess the relationship between upper respiratory tract SARS-CoV-2 IgG levels in fully vaccinated patients and the recovery of infectious virus on cell culture, ELISA was performed on upper respiratory samples from fully vaccinated individuals infected with Alpha (N = 43) or Delta (N = 24) variants as well as control unvaccinated but infected groups (Alpha, N = 30 and Delta, N = 17). A significant increase in localized IgG levels was observed in vaccinated versus unvaccinated individuals infected for the Alpha variant (Alpha unvaccinated, 0% positives, vaccinated 46.5% positives, p < 0.0001). More vaccinated individuals infected by the Delta variant showed detectable upper respiratory tract IgG but the mean IgG levels were not different between the groups (Delta unvaccinated 11.8% positives, vaccinated 37.5% positives) (Figure 4A). Vaccine breakthrough patients from both Alpha and Delta variants demonstrated an inverse correlation between upper respiratory tract IgG levels and the recovery of infectious virus on cell culture, regardless of Ct value (Figure 4 B and C).

**Figure 4.**
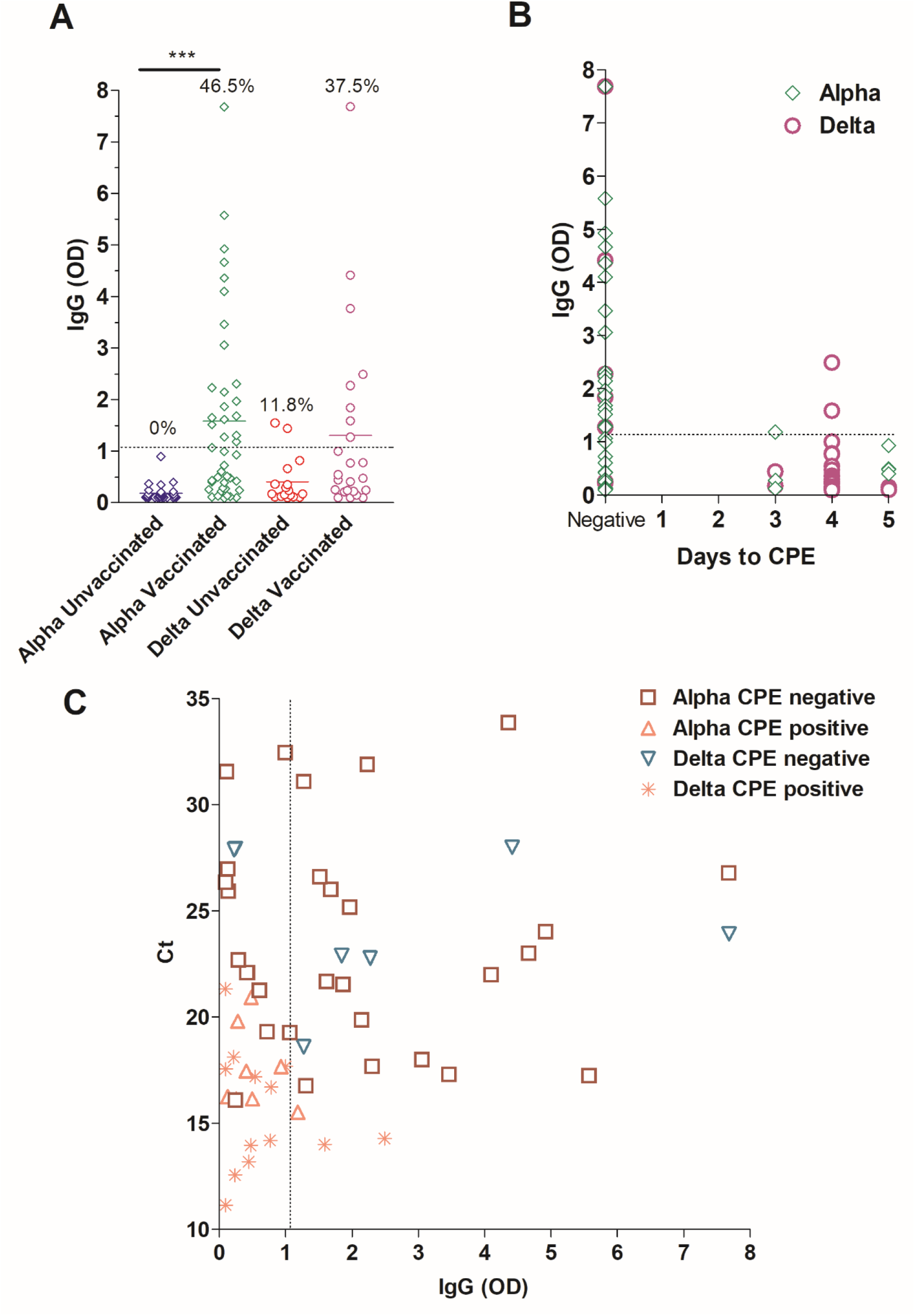
Local SARS-CoV-2 IgG in upper respiratory samples. A) IgG levels by ELISA in the upper respiratory samples collected from patients with Alpha or Delta variants (unvaccinated: Alpha, N = 30, Delta, N = 17, vaccinated: Alpha, N = 43, Delta, N = 24). One-way ANOVA *** p < 0.0001 B) SARS-CoV-2 IgG correlation to days to the first appearance of cytopathic effect (CPE) for Alpha and Delta true breakthrough specimens on Vero-TMPRSS2 cells. C) Correlation between local IgG levels, Ct values, and recovery of infectious virus. Dashed line demarcates the limit of borderline and negative ELISA results as specified per assay’s package insert. Define the p value and state the statistical tests used.

## Discussion

Alpha and Delta variants have raised major concerns associated with their evident success in displacing circulating variants and reaching global predominance. In the US, the Alpha variant predominated between February and June followed by the Delta variant that has become the most common lineage as of July, 2021 (https://covid.cdc.gov/covid-data-tracker/#variant-proportions). Our surveillance data collected from a wide geographical region in Washington DC, Virginia, and Baltimore showed an increase in the Alpha variant with a significant shift to a predominance in the second half of March (4) and in spite of detecting the Beta variant at the end of January (24). The Delta variant, on the other hand, was able to displace the Alpha and has been raising concerns associated with a notable increase in symptomatic breakthrough infections after full vaccination (10, 15) and its increased transmissibility. The virological, biological, and geographical determinants of the success of certain variants and their impact on the pandemic are not yet clearly understood. Different hypotheses including viral genomic changes associated with enhanced host cell receptor binding (25), higher viral loads (26), or escape from the neutralizing antibodies (27), have been proposed, but consistent evidence is lacking.

In this study, we analyzed a large cohort of samples diagnosed at Johns Hopkins clinical virology laboratory between January and July 2021 and compared the clinical presentations and disease outcomes in patients infected with the Delta versus the Alpha variants. Our data showed that the Alpha variant associated with increased hospitalization and ICU level care when compared to the previously predominant lineage, and infections with the Delta variant showed similar disease outcome to Alpha. It is of note though that our cohort infected with the Alpha variant was associated with higher comorbidities including diabetes and kidney disease compared to the Delta variant infections. As more Delta infected patients are identified in our laboratory and with more extended time for observing patients for outcomes, the clinical severity of the Delta variant will be better characterized.

The main observation notable in our cohort was the association of the Delta with more breakthrough infections and the significant increase in the days since receiving the vaccine in these cases. Breakthrough infections with the Delta were also associated with higher viral loads and increased recovery of infectious virus on cell culture when compared to breakthrough infections with the Alpha variant. We previously showed that vaccination was associated with reduction of the recovery of infectious virus on cell culture in a cohort that was primarily infected with the Alpha variant between January and May 2021, and this was associated with higher upper respiratory tract IgG levels (28). One possible explanation for the increased vaccine breakthrough infections seen with Delta is waning immune responses in vaccinated individuals as a result of the extended time post vaccination. We assessed localized IgG levels in upper respiratory tracts in Delta and Alpha breakthrough infections. Interestingly, the recovery of infectious virus was mainly notable in samples with negative or low upper respiratory tract IgG levels and this was more prominent with the Delta group, a correlation that was independent of the relative viral loads in the specimens. Our data is consistent with a recent observation from Vietnam that associated Delta breakthrough infections with lower levels of neutralizing antibodies induced by vaccination and a study from Wisconsin that showed infectious virus recovery from vaccinated patients (16, 17). This observation has important implications for infection control and supports the July, 2021 CDC guidelines of universal masking that includes vaccinated individuals to reduce viral transmission (https://www.cdc.gov/coronavirus/2019-ncov/vaccines/fully-vaccinated.html). Our data also suggest that increasing the upper respiratory tract IgG – perhaps through booster vaccinations – could help reduce transmission and symptomatic infections.

A major finding in our study is the increase in recovery of infectious virus for the Delta group in unvaccinated patients as well which did not correlate to differences in the relative viral loads in the samples. This is evidence of increased fitness of Delta perhaps driven by viral genetic determinants that enhance infectivity or replication. Changes within the spike protein of the Delta variant are thought to lead to enhanced binding to the host cell receptor (ACE2) and the S: P681R change in particular might increase the S protein cleavage efficiency allowing for more efficient entry (29). In addition, the S: L452R could contribute to the noticeable reduction in neutralization by serum antibodies and monoclonal antibodies (30-32). It is notable though that the Delta carries multiple other mutations within the spike and in other regions of the genome (Figure S1). Those changes might contribute to other mechanisms that could increase the virus stability, enhance its replication, or increase its ability to evade the immune responses as previously described for SARS-CoV (33). Additional studies are required to dissect the mechanistic impact of these changes and this work is currently in progress by our group.

The limitations of our study include the lower numbers of Delta variant specimens due to the lower positivity in the month of July, 2021, the infrequent specimens collected after 5 days of symptoms onset for the Delta vaccine breakthrough infections, and the relatively shorter time frame of Delta circulation that limited the clinical evaluation of some of the hospitalized patients. In addition, the phenotypes with cell culture experiments are usually dependent on the cell lines used, however Vero-TMPRSS-2 cells have been shown to enhance the isolation of SARS-CoV-2 (34). Moreover, the lack of serum and localized SARS-CoV-2 IgG data prior to infection for vaccine breakthrough cases in our cohort does not allow for the differentiation between waning immune responses and low initial responses to vaccines.

Taken together, we hypothesize that the notable increase in the time since receiving the vaccines combined with increased fitness of the Delta variant predisposes both vaccinated and unvaccinated individuals to symptomatic SARS-CoV-2 infections that are associated with high viral loads and transmission. Non-pharmaceutical interventions that include universal masking and social distancing to diminish transmission might be warranted to help control the summer 2021 Delta surge in the US. Booster vaccinations in groups at high risk of severe COVID-19 should be investigated to help reduce the burden of COVID-19 on the medical infrastructure.

## Data Availability

All data are available either through the manuscript or has been deposited in repositories such as GISAID

## Data sharing

Whole genome data were made available publicly and raw genomic data requests could be directed to HHM.

## Acknowledgement

This study was only possible with the unique efforts of the Johns Hopkins Clinical Microbiology Laboratory faculty and staff. HHM is supported by the HIV Prevention Trials Network (HPTN) sponsored by the National Institute of Allergy and Infectious Diseases (NIAID). Funding was provided by the Johns Hopkins Center of Excellence in Influenza Research and Surveillance (HHSN272201400007C), National Institute on Drug Abuse, National Institute of Mental Health, and Office of AIDS Research, of the NIH, DHHS (UM1 AI068613), the NIH RADx-Tech program (3U54HL143541-02S2), National Institute of Health RADx-UP initiative (Grant R01 DA045556-04S1), Centers for Disease Control (contract 75D30121C11061), the Johns Hopkins University President’s Fund Research Response, the Johns Hopkins department of Pathology, and the Maryland department of health. EK was supported by Centers for Disease Control and Prevention (CDC) MInD-Healthcare Program (Grant Number U01CK000589). The views expressed in this manuscript are those of the authors and do not necessarily represent the views of the National Institute of Biomedical Imaging and Bioengineering; the National Heart, Lung, and Blood Institute; the National Institutes of Health, or the U.S. Department of Health and Human Services.

---

**Figure S1.**
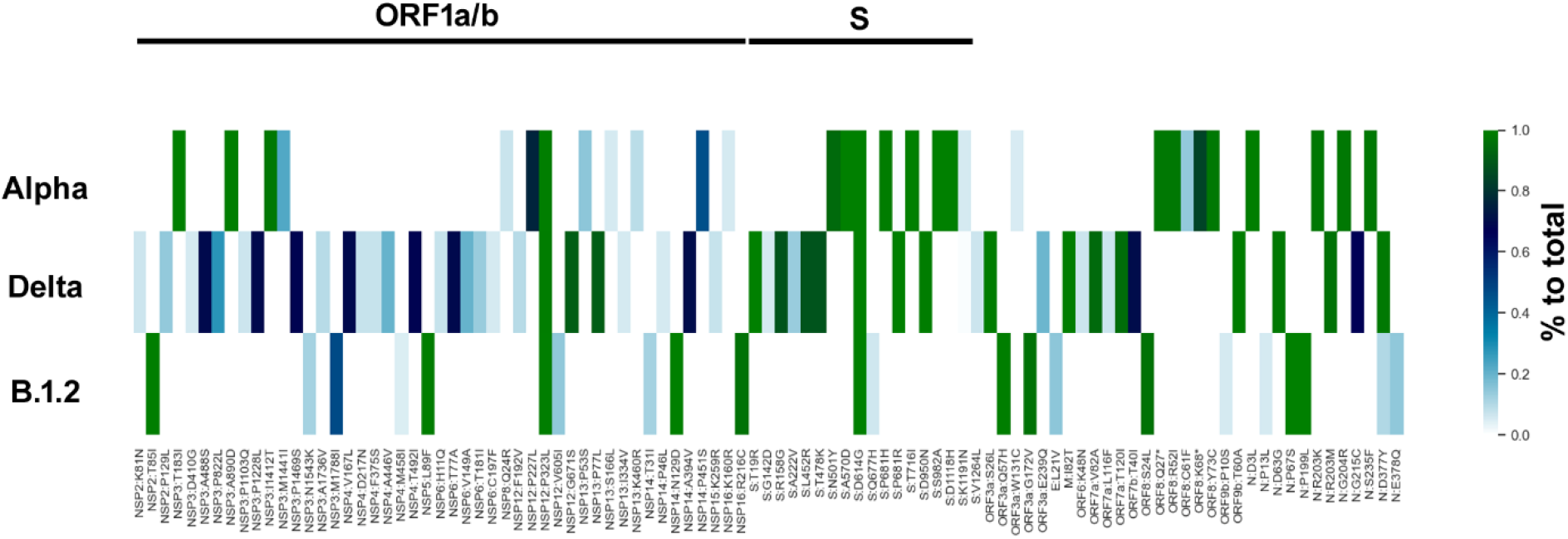
Common genomic amino acid changes in the Alpha versus Delta and B.1.2 variants. Included in the analysis are changes at more than 5% prevalence in sequenced samples for each variant.

## References

1. CDC. SARS-CoV-2 Variant Classifications and Definitions. (2021). Centers for Disease Control and Prevention. https://www.cdc.gov/coronavirus/2019-ncov/cases-updates/variant-surveillance/variant-info.html#Concern.

2. Public-Health-England. Investigation of novel SARS-CoV-2 variants of concern. (2021). https://www.gov.uk/government/publications/investigation-of-novel-sars-cov-2-variant-variant-of-concern-20201201.

3. Áine O’Toole VH, Oliver G. Pybus, Alexander Watts, Isaac I. Bogoch, Kamran Khan, Jane P. Messina, The COVID-19 Genomics UK (COG-UK) consortium, Network for Genomic Surveillance in South Africa (NGS-SA), Brazil-UK CADDE Genomic Network, Houriiyah Tegally, Richard R Lessells, Jennifer Giandhari, Sureshnee Pillay, Kefentse Arnold Tumedi, Gape Nyepetsi, Malebogo Kebabonye, Maitshwarelo Matsheka, Madisa Mine, Sima Tokajian, Hamad Hassan, Tamara Salloum, Georgi Merhi, Jad Koweyes, Jemma L Geoghegan, Joep de Ligt, Xiaoyun Ren, Matthew Storey, Nikki E Freed,Chitra Pattabiraman, Pramada Prasad, Anita S Desai, Ravi Vasanthapuram,Thomas F. Schulz, Lars Steinbrück, Tanja Stadler, Swiss Viollier Sequencing Consortium, Antonio Parisi, Angelica Bianco, Darío García de Viedma, Sergio Buenestado-Serrano, Vítor Borges, Joana Isidro, Sílvia Duarte, João Paulo Gomes, Neta S. Zuckerman, Michal Mandelboim, Orna Mor, Torsten Seemann, Alicia Arnott, Jenny Draper, Mailie Gall, William Rawlinson, Ira Deveson, Sanmarié Schlebusch, Jamie McMahon, Lex Leong, Chuan Kok Lim,Maria Chironna, Daniela Laconsole, Antonin Bal, Laurence Josset, Edward Holmes, Kirsten St George, Erica Lasek-Nesselquist, Reina S. Sikkema, Bas B. Oude Munnink, Marion Koopmans, Mia Brytting, V. Sudha Rani, S. Pavani, Teemu Smura, Albert Heim, Satu Kurkela, Massab Umair, Muhammad Salman, Barbara Bartolini, Martina Rueca, Christian Drosten, Thorsten Wolff, Olin Silander, Dirk Eggink, Chantal Reusken, Harry Vennema, Aekyung Park, SEARCH Alliance San Diego, National Virus Reference Laboratory, SeqCOVID-Spain, Danish Covid-19 Genome Consortium (DCGC), Communicable Diseases Genomic Network (CDGN), Dutch National SARS-CoV-2 surveillance program,#, Division of Emerging Infectious Diseases KDCA, Tulio de Oliveira, Nuno R. Faria, Andrew Rambaut, Moritz U. G. Kraemer. Tracking the international spread of SARS-CoV-2 lineages B.1.1.7 and B.1.351/501Y-V2. (2021) https://virological.org/t/tracking-the-international-spread-of-sars-cov-2-lineages-b-1-1-7-and-b-1-351-501y-v2/592.

4. Morris CP, Luo CH, Amadi A, Schwartz M, Gallagher N, Ray SC, Pekosz A, Mostafa HH. 2021. An Update on SARS-CoV-2 Diversity in the United States National Capital Region: Evolution of Novel and Variants of Concern. Clin Infect Dis doi:10.1093/cid/ciab636.

5. Davies NG, Abbott S, Barnard RC, Jarvis CI, Kucharski AJ, Munday JD, Pearson CAB, Russell TW, Tully DC, Washburne AD, Wenseleers T, Gimma A, Waites W, Wong KLM, van Zandvoort K, Silverman JD, Diaz-Ordaz K, Keogh R, Eggo RM, Funk S, Jit M, Atkins KE, Edmunds WJ. 2021. Estimated transmissibility and impact of SARS-CoV-2 lineage B.1.1.7 in England. Science 372:eabg3055.

6. Davies NG, Jarvis CI, van Zandvoort K, Clifford S, Sun FY, Funk S, Medley G, Jafari Y, Meakin SR, Lowe R, Quaife M, Waterlow NR, Eggo RM, Lei J, Koltai M, Krauer F, Tully DC, Munday JD, Showering A, Foss AM, Prem K, Flasche S, Kucharski AJ, Abbott S, Quilty BJ, Jombart T, Rosello A, Knight GM, Jit M, Liu Y, Williams J, Hellewell J, O’Reilly K, Chan Y-WD, Russell TW, Procter SR, Endo A, Nightingale ES, Bosse NI, Villabona-Arenas CJ, Sandmann FG, Gimma A, Abbas K, Waites W, Atkins KE, Barnard RC, Klepac P, Gibbs HP, Pearson CAB, Brady O, et al. 2021. Increased mortality in community-tested cases of SARS-CoV-2 lineage B.1.1.7. Nature doi:10.1038/s41586-021-03426-1.

7. Frampton D, Rampling T, Cross A, Bailey H, Heaney J, Byott M, Scott R, Sconza R, Price J, Margaritis M, Bergstrom M, Spyer MJ, Miralhes PB, Grant P, Kirk S, Valerio C, Mangera Z, Prabhahar T, Moreno-Cuesta J, Arulkumaran N, Singer M, Shin GY, Sanchez E, Paraskevopoulou SM, Pillay D, McKendry RA, Mirfenderesky M, Houlihan CF, Nastouli E. 2021. Genomic characteristics and clinical effect of the emergent SARS-CoV-2 B.1.1.7 lineage in London, UK: a whole-genome sequencing and hospital-based cohort study. Lancet Infect Dis doi:10.1016/S1473-3099(21)00170-5.

8. Chia PY, Xiang Ong SW, Chiew CJ, Ang LW, Chavatte J-M, Mak T-M, Cui L, Kalimuddin S, Chia WN, Tan CW, Ann Chai LY, Tan SY, Zheng S, Pin Lin RT, Wang L, Leo Y-S, Lee VJ, Lye DC, Young BE. 2021. Virological and serological kinetics of SARS-CoV-2 Delta variant vaccine-breakthrough infections: a multi-center cohort study. medRxiv doi:10.1101/2021.07.28.21261295:2021.07.28.21261295.

9. Brown CM, Vostok J, Johnson H, Burns M, Gharpure R, Sami S, Sabo RT, Hall N, Foreman A, Schubert PL, Gallagher GR, Fink T, Madoff LC, Gabriel SB, MacInnis B, Park DJ, Siddle KJ, Harik V, Arvidson D, Brock-Fisher T, Dunn M, Kearns A, Laney AS. 2021. Outbreak of SARS-CoV-2 Infections, Including COVID-19 Vaccine Breakthrough Infections, Associated with Large Public Gatherings - Barnstable County, Massachusetts, July 2021. MMWR Morb Mortal Wkly Rep 70:1059–1062.

10. Musser JM, Christensen PA, Olsen RJ, Long SW, Subedi S, Davis JJ, Hodjat P, Walley DR, Kinskey JC, Gollihar J. 2021. Delta variants of SARS-CoV-2 cause significantly increased vaccine breakthrough COVID-19 cases in Houston, Texas. medRxiv doi:10.1101/2021.07.19.21260808:2021.07.19.21260808.

11. Sheikh A, McMenamin J, Taylor B, Robertson C, Public Health S, the EIIC. 2021. SARS-CoV-2 Delta VOC in Scotland: demographics, risk of hospital admission, and vaccine effectiveness. Lancet 397:2461–2462.

12. Lopez Bernal J, Andrews N, Gower C, Gallagher E, Simmons R, Thelwall S, Stowe J, Tessier E, Groves N, Dabrera G, Myers R, Campbell CNJ, Amirthalingam G, Edmunds M, Zambon M, Brown KE, Hopkins S, Chand M, Ramsay M. 2021. Effectiveness of Covid-19 Vaccines against the B.1.617.2 (Delta) Variant. N Engl J Med doi:10.1056/NEJMoa2108891.

13. Thompson MG, Burgess JL, Naleway AL, Tyner H, Yoon SK, Meece J, Olsho LEW, Caban-Martinez AJ, Fowlkes AL, Lutrick K, Groom HC, Dunnigan K, Odean MJ, Hegmann K, Stefanski E, Edwards LJ, Schaefer-Solle N, Grant L, Ellingson K, Kuntz JL, Zunie T, Thiese MS, Ivacic L, Wesley MG, Mayo Lamberte J, Sun X, Smith ME, Phillips AL, Groover KD, Yoo YM, Gerald J, Brown RT, Herring MK, Joseph G, Beitel S, Morrill TC, Mak J, Rivers P, Poe BP, Lynch B, Zhou Y, Zhang J, Kelleher A, Li Y, Dickerson M, Hanson E, Guenther K, Tong S, Bateman A, Reisdorf E, et al. 2021. Prevention and Attenuation of Covid-19 with the BNT162b2 and mRNA-1273 Vaccines. New England Journal of Medicine 385:320–329.

14. Teyssou E, Soulie C, Visseaux B, Lambert-Niclot S, Ferre V, Marot S, Jary A, Sayon S, Zafilaza K, Leducq V, Schnuriger A, Wirden M, Houhou-Fidouh N, Charpentier C, Morand-Joubert L, Burrel S, Descamps D, Calvez V, Geneviève Marcelin A. 2021. The 501Y.V2 SARS-CoV-2 variant has an intermediate viral load between the 501Y.V1 and the historical variants in nasopharyngeal samples from newly diagnosed COVID-19 patients. medRxiv doi:10.1101/2021.03.21.21253498:2021.03.21.21253498.

15. Petra M, Steven K, Mahesh Shanker D, Guido P, Bo M, Swapnil M, Charlie W, Thomas M, Isabella F, Rawlings D, Dami AC, Sujeet S, Rajesh P, Robin M, Meena D, Shantanu S, Kalaiarasan P, Radhakrishnan VS, Adam A, Niluka G, Jonathan B, Oscar C, Partha C, Priti D, Daniela C, Tom P, Dr Chand W, Neeraj G, Raju V, Meenakshi A, The Indian S-C-GC, Citiid-Nihr BioResource Covid-19 Collaboration AM, o Hyeon L, Wendy SB, Samir B, Seth F, Leo J, Partha R, Anurag A, Ravindra KG. 2021. Nature Portfolio doi:10.21203/rs.3.rs-637724/v1.

16. Riemersma KK, Grogan BE, Kita-Yarbro A, Halfmann P, Kocharian A, Florek KR, Westergaard R, Bateman A, Jeppson GE, Kawaoka Y, O’Connor DH, Friedrich TC, Grande KM. 2021. Shedding of Infectious SARS-CoV-2 Despite Vaccination when the Delta Variant is Prevalent - Wisconsin, July 2021. medRxiv doi:10.1101/2021.07.31.21261387:2021.07.31.21261387.

17. Chau NVV, Ngoc NM, Nguyet LA, Quan VM, Ny NTH, Khoa DB, Phong NT, Toan LM, Hong2 NTT, Tuyen NTK, Phat VV, Nhu LNT, Truc NHT, That BTT, Thao HP, Nguyen T, Thao P, Vuong VT, Tam TTT, Tai NT, Bao HT, Nhung HTK, Minh NTN, Tien NTM, Huy NC, Choisy M, Man DNH, Ty DTB, Anh NT, Uyen LTT, Tu TNH, Yen Lm, L. NTD, Hung M, Truong NT, Thanh TT, Thwaites G, Tan LV. 2021. Transmission of SARS-CoV-2 Delta variant among vaccinated healthcare workers, Vietnam. Preprints with THE LANCET https://papers.ssrn.com/sol3/papers.cfm?abstract_id=3897733.

18. Jarrett J, Uhteg K, Forman MS, Hanlon A, Vargas C, Carroll KC, Valsamakis A, Mostafa HH. 2021. Clinical performance of the GenMark Dx ePlex respiratory pathogen panels for upper and lower respiratory tract infections. J Clin Virol 135:104737.

19. Mostafa HH, Carroll KC, Hicken R, Berry GJ, Manji R, Smith E, Rakeman JL, Fowler RC, Leelawong M, Butler-Wu SM, Quintero D, Umali-Wilcox M, Kwiatkowski RW, Persing DH, Weir F, Loeffelholz MJ. 2020. Multi-center Evaluation of the Cepheid Xpert(R) Xpress SARS-CoV-2/Flu/RSV Test. J Clin Microbiol doi:10.1128/JCM.02955-20.

20. Mostafa HH, Hardick J, Morehead E, Miller JA, Gaydos CA, Manabe YC. 2020. Comparison of the analytical sensitivity of seven commonly used commercial SARS-CoV-2 automated molecular assays. J Clin Virol 130:104578.

21. Uhteg K, Jarrett J, Richards M, Howard C, Morehead E, Geahr M, Gluck L, Hanlon A, Ellis B, Kaur H, Simner P, Carroll KC, Mostafa HH. 2020. Comparing the analytical performance of three SARS-CoV-2 molecular diagnostic assays. J Clin Virol 127:104384.

22. Thielen PM, Wohl S, Mehoke T, Ramakrishnan S, Kirsche M, Falade-Nwulia O, Trovao NS, Ernlund A, Howser C, Sadowski N, Morris CP, Hopkins M, Schwartz M, Fan Y, Gniazdowski V, Lessler J, Sauer L, Schatz MC, Evans JD, Ray SC, Timp W, Mostafa HH. 2021. Genomic diversity of SARS-CoV-2 during early introduction into the Baltimore-Washington metropolitan area. JCI Insight 6.

23. Gniazdowski V, Morris CP, Wohl S, Mehoke T, Ramakrishnan S, Thielen P, Powell H, Smith B, Armstrong DT, Herrera M, Reifsnyder C, Sevdali M, Carroll KC, Pekosz A, Mostafa HH. 2020. Repeat COVID-19 Molecular Testing: Correlation of SARS-CoV-2 Culture with Molecular Assays and Cycle Thresholds. Clin Infect Dis doi:10.1093/cid/ciaa1616.

24. Feder KA, Pearlowitz M, Goode A, Duwell M, Williams TW, Chen-Carrington PA, Patel A, Dominguez C, Keller EN, Klein L, Rivera-Colon A, Mostafa HH, Morris CP, Patel N, Schauer AM, Myers R, Blythe D, Feldman KA. 2021. Linked Clusters of SARS-CoV-2 Variant B.1.351 - Maryland, January-February 2021. MMWR Morb Mortal Wkly Rep 70:627–631.

25. Muik A, Wallisch A-K, Sänger B, Swanson KA, Mühl J, Chen W, Cai H, Maurus D, Sarkar R, Türeci Ö, Dormitzer PR, Şahin U. 2021. Neutralization of SARS-CoV-2 lineage B.1.1.7 pseudovirus by BNT162b2 vaccine–elicited human sera. Science 371:1152–1153.

26. Kidd M, Richter A, Best A, Cumley N, Mirza J, Percival B, Mayhew M, Megram O, Ashford F, White T, Moles-Garcia E, Crawford L, Bosworth A, Atabani SF, Plant T, McNally A. 2021. S-variant SARS-CoV-2 lineage B1.1.7 is associated with significantly higher viral loads in samples tested by ThermoFisher TaqPath RT-qPCR. The Journal of Infectious Diseases doi:10.1093/infdis/jiab082.

27. Diamond M, Chen R, Xie X, Case J, Zhang X, VanBlargan L, Liu Y, Liu J, Errico J, Winkler E, Suryadevara N, Tahan S, Turner J, Kim W, Schmitz A, Thapa M, Wang D, Boon A, Pinto D, Presti R, O’Halloran J, Kim A, Deepak P, Fremont D, Corti D, Virgin H, Crowe J, Droit L, Ellebedy A, Shi PY, Gilchuk P. 2021. SARS-CoV-2 variants show resistance to neutralization by many monoclonal and serum-derived polyclonal antibodies. Res Sq doi:10.21203/rs.3.rs-228079/v1.

28. Mostafa HH, Luo CH, Morris CP, Li M, Swanson NJ, Amadi A, Gallagher N, Pekosz A. 2021. SARS-CoV-2 Infections in mRNA Vaccinated Individuals are Biased for Viruses Encoding Spike E484K and Associated with Reduced Infectious Virus Loads that Correlate with Respiratory Antiviral IgG levels. medRxiv doi:10.1101/2021.07.05.21259105:2021.07.05.21259105.

29. Scudellari M. 2021. How the coronavirus infects cells - and why Delta is so dangerous. Nature 595:640–644.

30. Lucas C, Vogels CBF, Yildirim I, Rothman JE, Lu P, Monteiro V, Gelhausen JR, Campbell M, Silva J, Tabachikova A, Muenker MC, Breban MI, Fauver JR, Mohanty S, Huang J, Initiative YS-C-GS, Pearson C, Muyombwe A, Downing R, Razeq J, Petrone M, Ott I, Watkins A, Kalinich C, Alpert T, Brito A, Earnest R, Murphy S, Neal C, Laszlo E, Altajar A, Tikhonova I, Castaldi C, Mane S, Bilguvar K, Kerantzas N, Ferguson D, Schulz W, Landry M, Peaper D, Shaw AC, Ko AI, Omer SB, Grubaugh ND, Iwasaki A. 2021. Impact of circulating SARS-CoV-2 variants on mRNA vaccine-induced immunity in uninfected and previously infected individuals. medRxiv doi:10.1101/2021.07.14.21260307:2021.07.14.21260307.

31. Planas D, Veyer D, Baidaliuk A, Staropoli I, Guivel-Benhassine F, Rajah MM, Planchais C, Porrot F, Robillard N, Puech J, Prot M, Gallais F, Gantner P, Velay A, Le Guen J, Kassis-Chikhani N, Edriss D, Belec L, Seve A, Courtellemont L, Pere H, Hocqueloux L, Fafi-Kremer S, Prazuck T, Mouquet H, Bruel T, Simon-Loriere E, Rey FA, Schwartz O. 2021. Reduced sensitivity of SARS-CoV-2 variant Delta to antibody neutralization. Nature 596:276–280.

32. Lazarevic I, Pravica V, Miljanovic D, Cupic M. 2021. Immune Evasion of SARS-CoV-2 Emerging Variants: What Have We Learnt So Far? Viruses 13.

33. Graham RL, Sparks JS, Eckerle LD, Sims AC, Denison MR. 2008. SARS coronavirus replicase proteins in pathogenesis. Virus Res 133:88–100.

34. Matsuyama S, Nao N, Shirato K, Kawase M, Saito S, Takayama I, Nagata N, Sekizuka T, Katoh H, Kato F, Sakata M, Tahara M, Kutsuna S, Ohmagari N, Kuroda M, Suzuki T, Kageyama T, Takeda M. 2020. Enhanced isolation of SARS-CoV-2 by TMPRSS2-expressing cells. Proceedings of the National Academy of Sciences 117:7001–7003.

